# Ovarian cancer recurrence prediction: comparing confirmatory to real world predictors with machine learning

**DOI:** 10.1101/2025.03.04.25321571

**Authors:** D. Katsimpokis, A.E.C. van Odenhoven, M.A.J.M. van Erp, H.H.B. Wenzel, M.A. van der Aa, M.M.H. van Swieten, H.P.M. Smedts, J.M.J. Piek

**Affiliations:** Department of Research & Development, Netherlands Comprehensive Cancer Organisation (IKNL), Utrecht, the Netherlands; Department of Gynaecology and Obstetrics and Catharina Cancer Institute, Catharina Hospital Eindhoven, the Netherlands; Department of Gynaecology and Obstetrics, Amphia Hospital Breda, the Netherlands

**Keywords:** Ovarian cancer, cancer recurrence, prediction modeling, machine learning, real world data

## Abstract

**Introduction:** Ovarian cancer is one of the deadliest cancers in women, with a 5-year survival rate of 17-28% in advanced stage (FIGO IIB-IV) disease and is often diagnosed at advanced stage. Machine learning (ML) has the potential to provide a better survival prognosis than traditional tools, and to shed further light on predictive factors. This study focuses on advanced stage ovarian cancer and contrasts expert-derived predictive factors with data-driven ones from the Netherlands Cancer Registry (NCR) to predict progression-free survival.

**Methods:** A Delphi questionnaire was conducted to identify fourteen predictive factors which were included in the final analysis. ML models (regularized Cox regression, Random Survival Forests and XGBoost) were used to compare the Delphi expert-based set of variables to a real-world data (RWD) variable set derived from the NCR. A traditional, non-regularized, Cox model was used as the benchmark.

**Results:** While regularized Cox regression models with the RWD variable set outperformed the traditional Cox regression with the Delphi variables (c-index: 0.70 vs. 0.64 respectively), the XGBoost model showed the best performance overall (c-index: 0.75). The most predictive factors for recurrence were treatment types and outcomes as well as socioeconomic status, which were not identified as such by the Delphi questionnaire.

**Conclusion:** Our results highlight that ML algorithms have higher predictive power compared to the traditional Cox regression. Moreover, RWD from a cancer registry identified more predictive variables than a panel of experts. Overall, these results have important implications for AI-assisted clinical prognosis and provide insight into the differences between AI-driven and expert-based decision-making in survival prediction.

## 1. Introduction

Ovarian cancer is the fifth deadliest cancer in women worldwide and has the highest mortality rate of all gynecological cancers [1]. In the Netherlands, approximately 1300 women are diagnosed with ovarian cancer annually, of whom around 1000 will die from the disease, which is usually diagnosed at a late stage [2]. The five-year survival rate is only 17-28% for women with advanced stage (Federation of International Gynaecology and Obstetrics [FIGO] classification stages IIB-IV) [3, 4]. Even after initial treatment, approximately 70% to 80% of patients develop recurrent disease [5].

Despite decades of research on risk factors for ovarian cancer recurrence, such as FIGO classification and tumor characteristics, it is still challenging to accurately predict progression-free survival for (individual) ovarian cancer patients [6–8]. In recent years, great promise has come from the field of artificial intelligence (AI) and machine learning (ML), where evidence for timely detection and accurate cancer prognosis has grown tremendously [9]. By learning statistical associations from large amounts of data, ML models can show high accuracy of diagnostic prediction, sometimes comparable to or better than health-care professionals (e.g., by spotting cancerous masses in magnetic resonance imaging data) and time-to-event modeling (e.g., overall/progression-free survival) thereby paving the way towards AI-assisted healthcare [10, 11].

Specifically in the field of gynecological cancers, the use of AI has also gained ground. [12] For example, a recent meta-analysis of 34 imaging studies using AI to detect ovarian cancer across different imaging modalities, showed that algorithms had equal and, sometimes, higher diagnostic accuracy than clinicians [13]. ML models have also been developed to differentiate benign from malignant ovarian neoplasms, to predict overall survival after diagnosis, and to identify biomarkers and demographic factors contributing to overall survival [14–16]. Most studies related to AI and ML in ovarian cancer have focused on the histopathology, diagnosis and performance. However, prediction of progression-free survival and its related predictive factors has received less attention [12].

The current study aims to identify predictive factors for ovarian cancer recurrence and evaluate the performance of ML models in comparison to traditional models, such as Cox regression. We specifically focused on patients with advanced-stage disease who face the most challenging survival profile. We featured two sets of predictive variables in our analysis: one set selected by medical experts through a Delphi consensus process, and a larger set derived from real world data (RWD) obtained from the Netherlands Cancer Registry (NCR). This work has important implications for AI-assisted clinical prognosis and provides insight into the differences between AI and medical expert-based variable selection for survival prediction.

## 2. Data and Methods

### 2.1. Data source

We extracted data on ovarian cancer from the NCR, which is the national population-based cancer registry of the Netherlands, with nationwide coverage since 1989. The registry offers cancer incidence information, along with clinical, socio-demographic and (primary) treatment data. Information on vital status and date of death is updated annually using an automated link with the national Personal Records Database (BRP), and is now up to date until February 1st, 2024.

### 2.2. Data sample

Our initial sample consisted of patients diagnosed with epithelial ovarian cancer in the Netherlands from 2015 – 2017 (n = 3667). We excluded 63 patients with incomplete vital status information. We focused on advanced stages (FIGO stage: IIB – IV; although some where clinically suspect of having early stage disease), because these patients have a much higher mortality rate than those with early stage disease, thereby further excluding 731 patients. Subsequently, we restricted the sample further in order to avoid two common biases. Firstly, we excluded 489 patients that did not receive any therapies, as it would act as a competing risk and, therefore, bias cumulative risk estimators [17]. In addition, our sample included information on (the timing of) various types of therapy (i.e. neoadjuvant, primary and adjuvant; primary and interval debulking surgery). Since some therapies can take longer to complete than others, they would present inflated survivorship estimates in comparison to the shorter therapy regimes only because people need to survive long enough to finish them, leading to immortal time bias [18]. Our data showed evidence of immortal time bias (see Appendix A). Since the vast majority (>85%) of the patients had completed their primary therapy within the first 5 months after diagnosis, we excluded a further 355 patients who had disease recurrence before the first 5 months and we started following patients from 5 months after diagnosis. After exclusions, the final sample consisted of 2029 patients. A flow-chart can be found in Figure 1.

**Figure 1.**
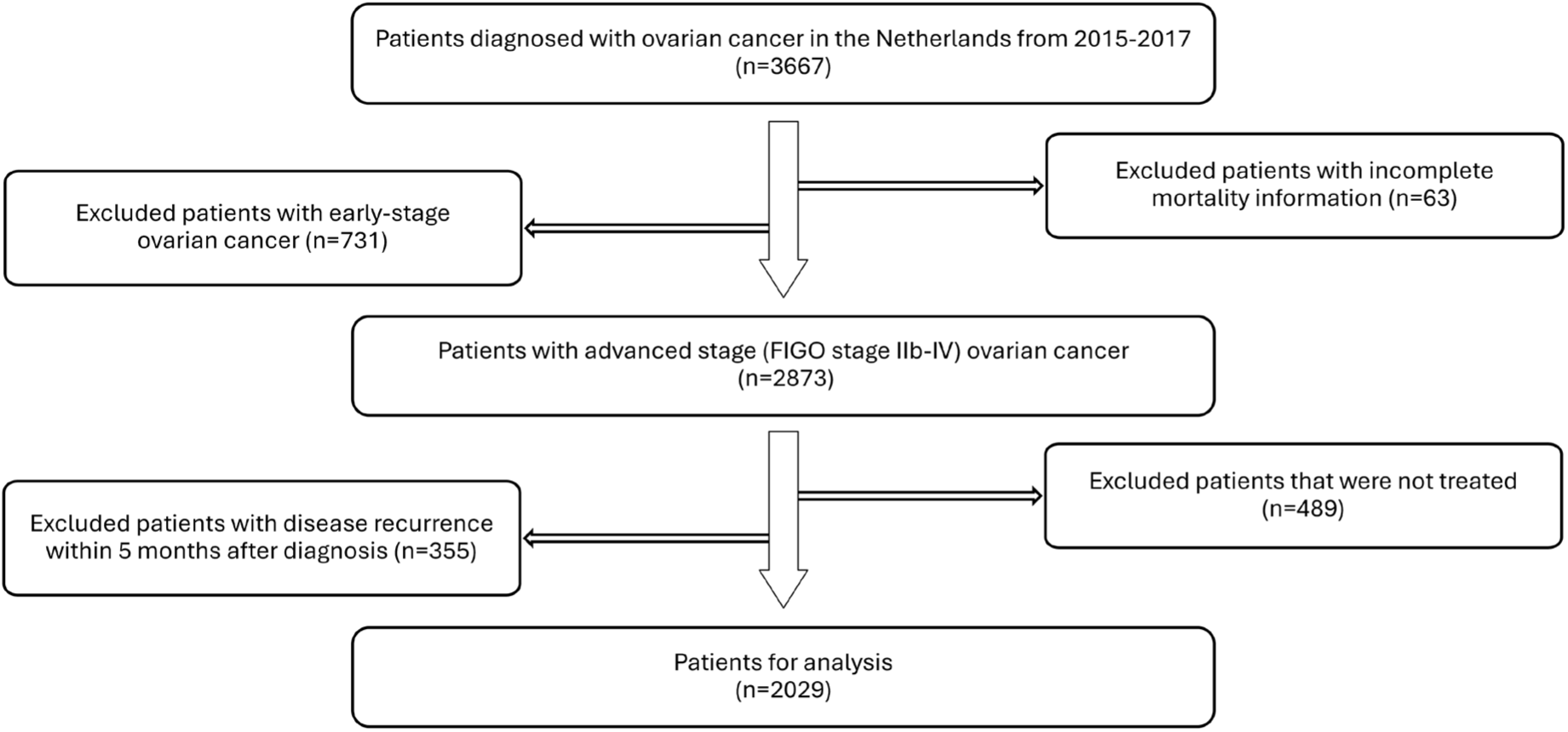
Flow chart of patient selection.

### 2.3. Delphi consensus making

A Delphi questionnaire was conducted to reach expert consensus [19]. The expert panel consisted of 40 medical doctors (oncologists, gynecologic oncologists, surgical oncologists), 20 of which ended up filling in the questionnaires. We asked participants to answer the following question: “in your experience as a medical specialist, what factors play a role in survival and disease free survival in patients with high stage (FIGO IIb-IV) ovarian cancer?” Out of all responces, we identified 20 possible prognostic factors for recurrent disease. The information for six factors was not available in the NCR. Thus, fourteen expert-identified prognostic factors were included in the analysis (see Table 1). Detailed information about the full Delphi process and the outcomes of the questionnaire are presented in the supplementary material. All data were evaluated and analysed by two clinical investigators (AvO, MvE).

**Table 1.** Delphi variables agreed by experts as predictive factors, presented in alphabetical order. The second column indicates the presence of information in the NCR. The last column lists NCR variables that encodes this information. “✔” shows presence in the NCR, while “–” shows absence or practically absence (more than 90%) of information in the NCR. Abbreviations: PCI: Peritoneal Cancer Index. CA-125: Cancer Antigen 125, ASA: American Society of Anesthesiologists. CCI: Charlston Comorbidity Index score.

| **Delphi-suggested variables** | **Presence in the NCR** | **NCR-related variables** |
| --- | --- | --- |
| *Age of the patient* | ✓ | Age at diagnosis |
| *Ascites* | ✓ | – |
| *CA-125 after chemotherapy* | – | – |
| *Chemotherapy-surgery in-between time* | ✓ | Chemotherapy-surgery in-between time |
| *Comorbidities* | ✓ | CCI |
| *Condition of the patient* | ✓ | Performance Status, ASA, CCI |
| *Dosage chemotherapy complete* | ✓ | Number of platinum cycles |
| *FIGO classification* | ✓ | FIGO classification |
| *Genetic predisposition* | ✓ | BRCA molecular diagnostics |
| *Histological subtype of the tumor* | ✓ | Histological subtype |
| *Involvement of mesentery* | – | – |
| *Level of PCI* | – | – |
| *Motivation of the patient* | – | – |
| *Nutritional status* | ✓ | BMI |
| *Pattern of metastases (lymph nodes involved)* | ✓ | Number of positive lymph nodes |
| *Quality of surgical team* | – | – |
| *Response on chemotherapy* | – | – |
| *Time to recurrence* | ✓ | Time to recurrence |
| *Tumor biology* | ✓ | BRCA molecular diagnostics |
| *WHO performance status* | ✓ | Performance status |

### 2.4. Data variables

#### 2.4.1. Delphi approach

This set includes fourteen predictor variables from the Delphi consensus. These variables constitute the theoretical (i.e., expert-based) minimum number of predictors for progression-free ovarian cancer survival. These variables were defined without seeing the data, thereby constituting a confirmatory test [20].

#### 2.4.2. Data-driven approach

For the data-driven approach, we relied on a larger RWD set of prognostic factors. In this case, the following NCR variables were included on top of the Delphi variables: *pretreatment CA-125, ovariectomy (no vs. yes), ovariectomy with hysterectomy (no vs. yes), staging surgery (no vs. yes), result of debulking (complete vs. optimal vs. suboptimal vs. no debulking), and type of debulking (interval vs. primary vs. no debulking), type of chemotherapy (preoperative vs. postoperative vs. pre&postoperative vs. no chemotherapy vs. yes, but no surgery), hormonal therapy (no vs. yes), intraperitoneal chemotherapy (no vs. yes), hyperthermic intraperitoneal chemotherapy (no vs. yes), differentiation grade (levels: 1-3), laterality (bilateral vs. unilateral vs. extra-ovarian), socio-economic status (levels: 1-10)*.

### 2.5. Machine learning modeling

#### 2.5.1. Cox-based models

For the Delphi approach, we used the semi-parametric Cox proportional hazards model with the Delphi variables. This makes the model easy to interpret because it remains agnostic about the baseline hazard function while treating the predictors in a regression-type form. However, the model relies on proportional hazards assumptions, which can be restrictive and may not always hold true in practice [21].

In our RWD approach, we applied two adaptations of the Cox model with the full set of variables: (i) the Cox model with the Lasso regularization, and (ii) the Cox model with the Elastic Net, which combines Lasso and Ridge Regularizations. Regularization is crucial for two reasons. First, it has been shown to enhance the performance of the Cox model [22, 23]. Second, it prevents overfitting by shrinking non-predictive coefficients to zero, effectively performing variable selection. However, compared to Lasso, Elastic Net tends to perform more stable variable selection in the presence of correlated variables [24]. Therefore, we included both models in subsequent analyses for comparison.

#### 2.5.2. Model fitting

The full model fitting procedure is described in Appendix B. In short, missing values were imputed using the Missforest algorithm [25]. The models were fitted using internal validation based on bootstrapping (n = 500) to adjust for generalization error [26]. Model parameters were optimized during the internal validation step using a randomized cross-validation parameter search. Bootstrapping was used to achieve stable variable selection estimates.

#### 2.5.3. More advanced machine learning modeling

While Cox-based models are easily interpretable, we also explored Random Survival Forests (RSFs) and the extreme gradient boosting trees model (XGBoost), which have demonstrated state-of-the-art performance in order to evaluate their marginal predictive advantage within the RWD approach [27–30]. Both RSFs and XGBoost are not restricted by the proportional hazards assumption, allowing them to capture non-linear relationships in the data. Moreover, since XGBoost can natively handle missing values, we fitted imputed and non-imputed versions of the RWD to this model to investigate the impact of imputation on performance. Appendix B provides more details.

## 3. Results

Table 2 gives the descriptive statistics of the cohort (i.e. variables of the Delphi and the data-driven approach). 77% of patients (i.e., no recurrence = 467; recurrence = 1562) showed disease recurrence after the start of follow-up, which began 5 months after the initial diagnosis, with mean time-to-recurrence at 600 days (standard deviation: 570 days) with a maximum follow-up time of 2043 days.

**Table 2.** Descriptive statistics of predictor variables (in alphabetical order). In case of categorical or ordinal variables, the count and percentage of each level are given. In case of numerical variables the mean and standard deviation (“SD”) are given. The additional variables of the RWD approach are indicated with a star (*) in the first column. Abbreviations respectively: SD: Standard deviation, ASA: American Society of Anesthesiologists, BMI: Body Mass Index, NA: Not applicable, BRCA: BReast CAncer gene, CCI: Charlson Comorbidity Index, FIGO: Federation of International Gynaecology and Obstetrics, WHO: World Health Organisation, NOS: Not Otherwise Specified, HIPEC: Hyperthermic intraperitoneal chemotherapy, CA-125: Cancer Antigen-125, SES: socio economic status.

|  |  |  |
| --- | --- | --- |
| **Age of the patient** (in years) - mean [SD*] |  | 66 [10.8] |
| **ASA-classification** (n (%)) | I<br>II<br>III<br>NA* | 295 (14)<br>1081 (53)<br>306 (15)<br>347 (17) |
| **BMI** *(in kg/m3) - mean [SD] |  | 25.8 [4.76] |
| **BRCA\* mutation** – n (%) | BRCA 1<br>BRCA 2<br>No BRCA mutation<br>No BRCA mutation analysis<br>NA | 106 (5)<br>66 (3)<br>1056 (53)<br>336 (17)<br>465 (23) |
| **CCI\*** – n (%) | 0<br>1<br>2 (two or more)<br>NA | 1246 (61)<br>467 (23)<br>149 (7)<br>167 (8) |
| **Differentiation grade** – n (%) | 1<br>2<br>3<br>NA | 141 (7)<br>86 (4)<br>1352 (67)<br>450 (22) |
| **Chemotherapy** – n (%) | No<br>Only postoperative<br>Pre-and post-operative<br>Only pre-operative<br>Yes, but no surgery | 76 (4)<br>584 (29)<br>1019 (50)<br>39 (19)<br>311 (15) |
| **Time between chemotherapy and surgery**<br>(in days) – mean [SD] |  | 42.7 [44.8] |
| **Debulking result** – n (%) | Complete<br>Optimal<br>Incomplete<br>No debulking<br>NA | 1046 (52)<br>428 (21)<br>121 (6)<br>416 (21)<br>18 (0.8) |
| **Debulking type** – n (%) | Primary<br>Interval<br>No debulking<br>NA | 565 (28)<br>1048 (52)<br>416 (21)<br>18 (0.8) |
| **FIGO\* classification** – n (%) | IIB<br>III<br>IV | 209 (10)<br>1254 (62)<br>566 (28) |
| **WHO\* performance status** – n (%) | 0<br>1<br>2<br>3<br>NA | 893 (43)<br>459 (23)<br>108 (5)<br>27 (1)<br>542 (27) |
| **Histological subtype of the tumor** – n (%) | Adenocarcinoma NOS*<br>Clear cell<br>Endometrioid | 173 (8)<br>83 (4)<br>53 (3) |
|  | *Mucinous* | 41 (2) |
|  | *Other* | 74 (4) |
|  | *Serous* | 1605 (79) |
| **Hormonal therapy – n (%)** | *No* | 1966 (97) |
|  | *Yes* | 63 (3) |
| **HIPEC\* – n (%)** | *No* | 2010 (99) |
|  | *Yes* | 19 (1) |
| **Ovariectomy with hysterectomy – n (%)** | *No* | 2010 (99) |
|  | *Yes* | 19 (1) |
| **Intraperitoneal chemotherapy – n (%)** | *No* | 1964 (97) |
|  | *Yes* | 65 (3) |
| **Staging surgery – n (%)** | *No* | 1978 (97) |
|  | *Yes* | 51 (3) |
| **Laterality – n (%)** | *Bilateral* | 827 (40) |
|  | *Other* | 201 (10) |
|  | *Unilateral* | 747 (39) |
|  | *NA* | 254 (12) |
| **Number of platinum based chemotherapy cycles – n (%)** | *6* | 1522 (75) |
|  | *<6* | 288 (15) |
|  | *>6* | 219 (11) |
| **Ovariectomy – n (%)** | *No* | 1962 (97) |
|  | *Yes* | 67 (3) |
| **Pretreatment CA-125 – mean [SD]** |  | 1613.7 [31113.1] |
| **SES\*(deciles) – n (%)** | *1* | 217 (11) |
|  | *2* | 211 (10) |
|  | *3* | 234 (12) |
|  | *4* | 221 (11) |
|  | *5* | 227 (11) |
|  | *6* | 200 (10) |
|  | *7* | 191 (9) |
|  | *8* | 182 (9) |
|  | *9* | 173 (9) |
|  | *10* | 164 (8) |
|  | *NA* | 9 (<1) |

The comparison of the three Cox models indicated that the baseline (Delphi) model revealed lower predictive accuracy than the RWD approach (cf. Table 3). This shows that the additional cancer registry variables have an added predictive value beyond the Delphi variables. Conversely, some predictors identified in the Delphi process may not be very predictive in the presence of the RWD variable set. Although both regularization models achieved the same performance, we picked the Elastic Net model for variable selection, because it produces more stable coefficients than the Lasso model in the presence of correlated variables. Notably, calibration of the Elastic Net model indicated no gross misfit (see Appendix C).

**Table 3.** Model performance adjusted for generalization error (i.e., overfitting) based on bootstrapping (500 iterations). Mean and 95% quantile-based confidence intervals are given. Numbers indicate the concordance (c-)index. “NAs” indicate whether the data set contained missing values or whether these were imputed beforehand. Note that only the Cox model (baseline) was fitted to the Delphi variables alone, while the rest of the models were fitted to the RWD variable set. The best performing model is indicated in bold.

| Model | Mean<br>performance | CI<sub>95%</sub> |
| --- | --- | --- |
| *Cox model (baseline)* | 0.643 | [0.657, 0.627] |
| *Cox model (Lasso)* | 0.703 | [0.689, 0.717] |
| *Cox model (Elastic Net)* | 0.703 | [0.689, 0.716] |
| ***XGBoost (without NAs)*** | **0.754** | **[0.738, 0.773]** |
| *XGBoost (with NAs)* | 0.739 | [0.723, 0.760] |
| *Random Forests* | 0.721 | [0.708, 0.735] |

In comparison to the data-driven Cox-based models, the XGBoost model, which was also fitted on the RWD, achieved higher performance than the Lasso and Elastic Net models, both in the version with imputed data and in the version without data imputation (cf. Table 3). Data imputation did not change the performance of XGBoost, implying that the MissForest imputation method did not decrease performance. It is noteworthy that the XGBoost model showed a substantial improvement of performance over the baseline Cox regression model with the Delphi variables. Finally, RSFs scored lower than the XGBoost model, but performed better in comparison to the (regularized) Cox regression models.

Variable selection showed a variety of results (cf. Figure 2). The top ten predictive variables showed a mix of variables grouped into three categories, relating to 1) intervention type and outcome (i.e., type of surgery and results of debulking, presence of postoperative chemotherapy, number of platinum cycles), 2) clinical characteristics (i.e., differentiation grade, FIGO stage, endometrioid cancer type, BRCA mutation) and 3) demographics (i.e., socio-economic status). Interestingly, only five of the top ten variables were identified by the Delphi panel, namely FIGO stage, endometrioid cancer type, BRCA2 or no BRCA mutation and number of platinum cycles, highlighting the importance of the data-driven variables. Moreover, two of the Delphi variables, e.g. the number of positive lymph nodes and time from chemotherapy to surgery, had low predictive power regarding disease recurrence. Lastly, other variables identified by the Delphi consensus, such as age and BMI, were found to be moderately predictive.

**Figure 2.**
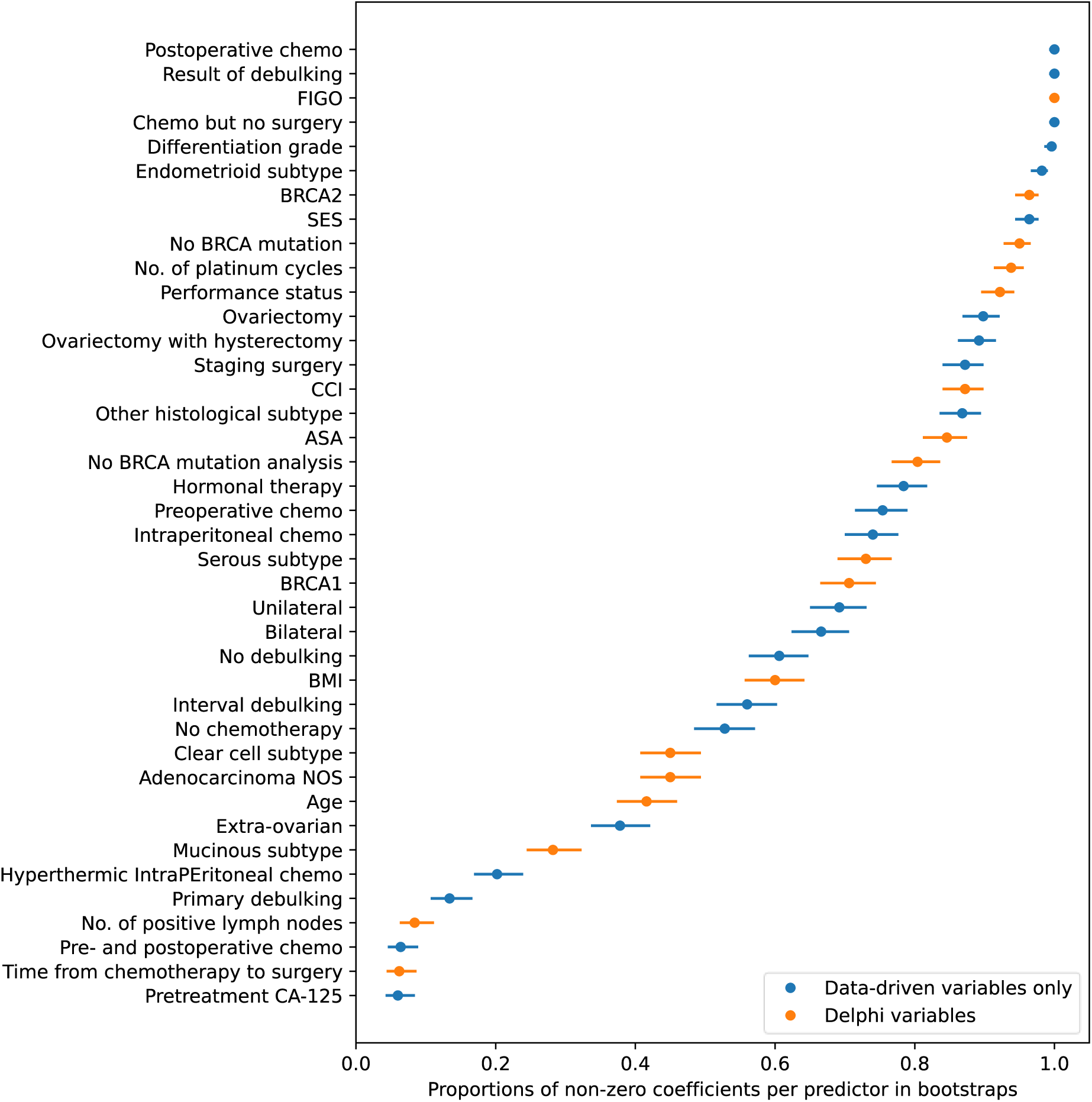
Variable selection from the Elastic Net model. Each dot indicates the proportion that each predictor was non-zero in the bootstrap samples, and the range indicates the estimated 95% Wilson-based confidence interval. The higher the percentage, the more important the variable is for prediction, and vice versa. Abbreviations: FIGO: Federation of International Gynaecology and Obstetrics, BRCA: BReast CAncer gene, CCI: Charlson Comorbidity Index, CCI: Charlson Comorbidity Index, ASA: American Society of Anesthesiologists Chemo: chemotherapy, NOS: not otherwise specified

## 4. Discussion

We applied and compared several machine learning models in order to predict progression-free survival of patients with ovarian cancer. We compared a confirmatory Delphi-based set of predictor variables with RWD drawn from the NCR that included the Delphi variables (i.e., nested comparison). Our results show that regularized Cox regression models based on the RWD variable set outperformed the traditional Cox regression based on the Delphi set (c-indices: 0.64 vs. 0.70 respectively). This means that the traditional Cox regression would correctly discriminate the time-to-event of a random pair of patients only 64% of the time, in contrast to the 70% of the regularized version based on the RWD set. Furthermore, variable selection on the regularized Cox model (i.e., Elastic Net) showed that many predictive variables for recurrence were related to types and results of therapy as well as socioeconomic status, all of which were not included in the Delphi set. Specifically, out of the ten most predictive variables, only five variables were identified by the Delphi consensus. Thirdly, the state-of-the-art XGBoost model achieved the best performance (c-index: 0.75) on the RWD variable list, outperforming both regularized Cox regression models and RSFs.

The superior model performance of the RWD approach underlines the importance of using data-driven, cancer registry variables for predicting ovarian cancer recurrence. Although the Delphi variables represent the (minimal) expert-based set of expected predictive factors for recurrence, not all of them were highly predictive (e.g., age), and some showed little to no predictive value (e.g., number of cancer-positive lymph nodes). Furthermore, many factors not mentioned in the Delphi consensus showed a high degree of predictiveness (e.g., socio-economic status). On the other hand, some variables that were not included in the Delphi consensus (e.g., pretreatment CA-125) played indeed a minimal role in recurrence prediction.

Our results indicate that while expert-based knowledge plays a valuable role in predicting ovarian cancer recurrence, it has limitations and should therefore be complemented with RWD from cancer registries. Future research can further enrich these data sets by incorporating primary healthcare data as well as more intricate demographic and time-varying information, to improve predictive accuracy. However, it is important to note that the predictive value of the factors analyzed in this study reflects the statistical correlations among these variables, rather than causal relationships. Establishing causality would require counterfactual intervention and a deeper understanding of the causal structure of the study system. [31] Such causal understanding, which is absent from ML models, may further explain why variable selection results from ML models and from the Delphi questionnaire did not completely overlap. This research question, which is beyond the scope of the current paper, could shed light on the medical decision-making during the Delphi process.

Variable selection through the Cox-based Elastic Net is easy to interpret. Due to their complexity, more advanced models such as RSFs and XGBoost require explainability methods to understand how the models use the predictor variables to perform prediction. Techniques such as Shapley values [32] can provide valuable insights into the contribution of each variable to the model’s prediction, but still require complex computations and are prone to overinterpretation (e.g., as causal indicators) or misinterpretation due to confirmation bias [33, 34]. In our current study, we opted to perform variable selection with the easily explainable Cox-based Elastic Net, since the performance of the XGBoost model was only marginally higher than the Cox-based models.

The superior performance of the XGBoost model is in line with recent literature suggesting that the model can outperform Cox-based models and other ensemble methods, such as RSFs, for time-to-event modeling [35]. Although the XGBoost technically outperformed the regularized Cox regression models, its advantage in predictive performance was not substantial. This result may stem from the fact that the proportional hazards assumption of the Cox model seems to not be grossly violated in our data (e.g., the calibration of the Cox models was good; cf. Appendix D). Higher sample sizes (i.e., many tens of thousands patient records) might be required for such non-linear patterns to be effectively used by XGBoost.

The current study has many methodological strengths. Firstly, the internal validation method based on bootstrapping validates the model building procedure and provides more stable performance metrics, including overfitting and variable selection, compared to simpler train-test split techniques [26]. Secondly, our sample size is larger than that in previous studies and we used a wide range of predictive factors, ranging from demographic, clinical and primary therapy information, thereby employing the strength of NCR information [36, 37]. Thirdly, we used an a priori expert-informed set of variables (i.e., the Delphi consensus) in order to have a theoretical baseline of the minimal set of predictive factors for recurrence disease before we see the data. This allowed us to separate a confirmatory test of this minimal set of variables from the expanded RWD set.

The study has also limitations. First of all, our model is only valid for prediction with follow-up starting at five months after diagnosis and for those patients that received at least some primary therapy, in order to avoid competing risk bias and immortal time bias, as the ML models employed cannot natively correct for these. Future research could employ more advanced methods, such as simulation to model competing risks. In addition, models such as time-varying-coefficient Cox regression could account for the different durations of the different combinations of primary therapies [38].

A second limitation of the current study concerns the variables suggested by the Delphi consensus but missing from the NCR (i.e., motivation of the patient, response on chemotherapy, level of PCI, CA-125 levels after chemotherapy, involvement of mesentery and quality of surgical team, ascites). In addition, some of the variables suggested by the Delphi (e.g., motivation of the patient or quality of surgical team) cannot be easily put down on a measurable scale. Future research could endeavor to find proxies for such variables, for example in patient self-reports on quality of life and external hospital reviews.

## 5. Conclusion

Our work provides evidence that ML algorithms, such as XGBoost, Random Survival Forests and regularized Cox regression are better in predicting progression-free survival for ovarian cancer than the traditional Cox model. In addition, expert-derived predictors can be enhanced through secondary (cancer registry) data, which can provide better predictive results when combined with ML variable selection.

## Data Availability

Data are not directly available, but need to be requested from Integraal Kankercentrum Nederland (IKNL) through a formal application procedure (www.iknl.nl)

## 6. Acknowledgements

DK thanks Maarten Bijlsma for his generous theoretical guidance throughout the project. Also thanks to all medical specialists’ who made the Delphi possible.

## 7. Statements and Declarations

### Ethics approval

According to the Dutch Central Committee on Research involving Human Subjects, no ethical approval is needed for this study, as it is a retrospective study, which uses data from the NCR.

